# Publishers’ responses to integrity concerns – does the type of notice influence subsequent citations? A cohort study

**DOI:** 10.64898/2026.02.25.26346683

**Authors:** Hugo Studd, Alison Avenell, Andrew Grey, Mark J Bolland

## Abstract

**Background:** Journals may respond to integrity concerns by publishing an editorial response (editorial notice, expression of concern (EoC) or retraction). We investigated whether the type of editorial response affected citation rates.

**Methods:** We obtained citations for 172 randomised controlled trials (RCTs) with integrity concerns (41 had editorial notices, 38 EoCs and 23 retractions) and control RCTs from the same journal and year. Monthly citation rates up to 60 months before and after editorial responses were compared by editorial response type, and to citation decline in control RCTs.

**Results:** 172 RCTs had 10,603 citations from 6,376 articles. 3,330 control trials were identified for 151/172 RCTs (15,948 citations, 87,811 articles). For both groups, citations increased steadily, peaking 45-65 months post-publication. There were no statistically significant differences in citation decline post-editorial response for trials receiving a retraction, EoC, or notice. Citations were lower in controls than index trials, so analyses were restricted to 1598 highly cited (>25) controls. The rate of decline for highly cited control trials was not statistically significantly different from the post-editorial response rate for index groups.

**Conclusion:** Citation rate decline after editorial responses did not differ by type of editorial response nor from the natural decline in control trials.

**Highlights:**

- Journals may respond to integrity concerns by issuing an editorial notice.
- The effect of expressions of concern or other editorial notices on citation patterns is unclear.
- Editorial notices did not accelerate citation decline compared with control trials.
- The type of notice was not associated with differences in citation decline.
- Late editorial notices appear ineffective in preventing continued citation.

## Introduction

Unfortunately, untrustworthy research gets published or, having been published, remains uncorrected or unretracted after integrity concerns are raised. The prevalence of unreliable publications is unknown but retractions are increasing in frequency,^1^ and surveys of researchers report that engagement in questionable research practices or misconduct is fairly common.^2-4^ Once integrity concerns about a publication have been notified to a journal or publisher, an investigation is usually conducted, with four possible outcomes: no action is taken, a correction is issued, an editorial response such as an expression of concern (EoC) is published, or the publication is retracted. More recently some publishers have begun to use editorial notices rather than EoCs to indicate that concerns have been raised about an article. A potentially important difference between an EoC and editorial notices is their visibility to readers: EoCs are more visible because they are indexed in bibliographic databases whereas editorial notices are only visible on journal websites. After the public notification of integrity concerns about an article, it would be expected that other authors would no longer cite the article because it is unreliable. Some analyses have concluded that this happens in practice, with retractions leading to decreases in citations,^5^ whereas others have reported persistence^6^ or even increased^7^ citation rates following retraction. We are not aware of research regarding the effect of other types of editorial responses on citation rates.

We wondered whether the type of editorial response - an editorial notice, an EoC, or a retraction-would affect the citation rates because of differences in their visibility or perceived seriousness. Previously, we simultaneously notified integrity concerns about a set of 172 randomised controlled trials (RCTs) published by a single research group^8^ that had accumulated 102 editorial responses at the time of these analyses, including 41 editorial notices, 38 EoC and 23 retractions. With this number and distribution of editorial responses, the dataset provides a chance to examine these hypotheses, specifically whether publication of any editorial response leads to a decline in citation rates that is greater than the decline seen in control publications without editorial responses, and secondly, whether the type of editorial response, EoC, editorial notice, or retraction had different effects on citations rates.

## Materials and Methods

In June 2025, we used the R package “citecorp” to obtain citations from the OpenCitations database (www.opencitations.net) for the previously identified 172 RCTs (the index set) from searches of Scopus, Web of Science and PubMed.^8^ We retrieved all citations using either the PMID, DOI or both for each article using the “oc_coci_cites” function. This retrieved an OpenCitations-specific “OMID” identifier for each citing article. We then used the “oc_coci_meta” function and this OMID identifier to extract the details for the citing article from the OpenCitations database. We limited citations to articles published before 2025 and excluded articles without a PMID/DOI if they were not categorised in OpenCitations as a journal article. If available, we used the PMID and/or DOI and the R “easyPubMed” package to obtain the date of publication (“pdat”) from the PubMed record. When both PMID and DOI were not available in OpenCitations (16% of citations), we were unable to use the PubMed publication date. Instead, we used the date of publication listed in OpenCitations. For 86 (0.8%) citations, no month of publication was reported, in which case we used June as the publication month.

To obtain a control group of RCTs, we searched for all RCTs published in the same journal during the same year as each publication in the index set. We searched PubMed using the easyPubMed package and the journal name, the term “randomized controlled trial [pt]” with a publication date in the relevant year. From the identified RCTs, we removed any publications listed in the Retraction Watch database, and any trials by the senior author of the index set of publications. For the control RCTs, we then followed the same approach as for the index RCTs. First, we retrieved all citations for each article from the OpenCitations database, then we extracted the details for the citing articles from the database, excluded articles as described previously, and then obtained the publication data from Pubmed (87%) or OpenCitations (13%, 1341 (1.1%) with no month).

Simple summary statistics were used to describe the two sets of publications. Next, we plotted mean citations per paper per month by the category of editorial response (EoCs, notices and retractions). We calculated the slope of the line of best fit for citations up to 60 months before or after the first editorial response for articles with an EoC, notice or retraction. For the index trials without any publisher action and the control RCTs, we used the “peak citation month”, the median of the months with the most citations as a reference point, to allow comparisons with the articles with editorial responses. For each time period, before or after the editorial response, the slopes for each category were compared to the slopes of the controls in a linear regression model and the interaction reported between time and category. For each individual paper, we calculated the slopes before and after the editorial response or peak citation month and compared them using a Wilcoxon test. We then calculated the differences between the slopes before and after the reference point and compared these differences between the various groups using a t-test because these differences were approximately normally distributed.

All analyses were undertaken using R 4.4.2 (R Core Team, 2024), and a p-value of <0.05 was considered statistically significant.

## Results

The 172 index RCTs^8^ were all published between Sept 2011 and January 2019. Figure 1 summarises the identification of control trials and citations. For the 172 index trials, there were 10,603 citations before 2025 from 6,376 unique articles. All 172 index trials were cited at least twice. The median number of citations per paper was 42.5, mean 62, and range 2-635. Of the 172 index trials, 70 had no editorial action and 102 had an editorial response: 41 received an editorial notice, 38 an EoC, and 23 a retraction.

**Figure 1.**
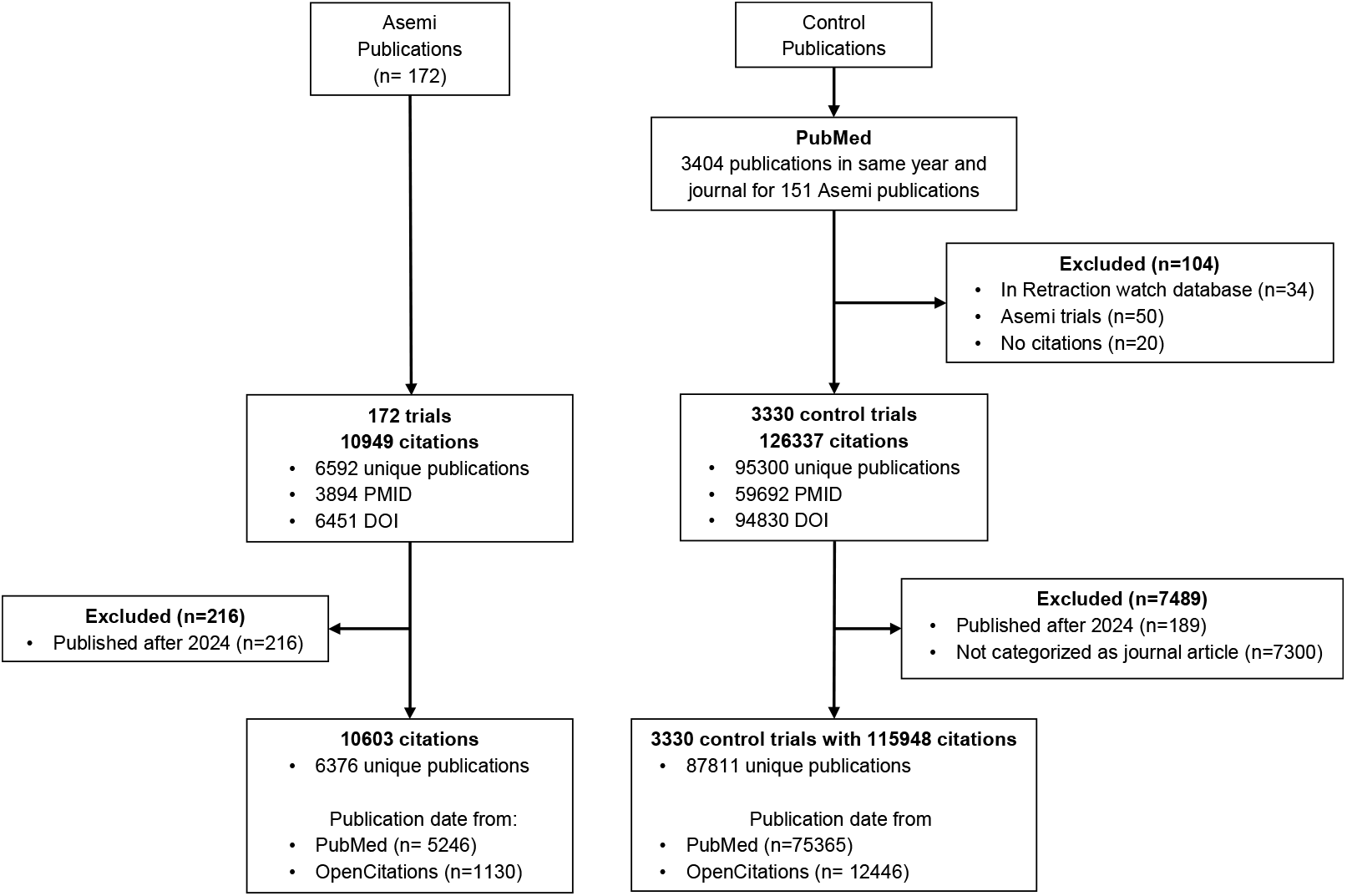
Flow sheet summarising the results of identifying control trials and citations.

We identified at least one RCT by other authors published in the same journal in the same year (“control trials”) for 151 index trials. The median number of controls per index trial was 13, mean 32, and range 1-253. Overall, there were 3,330 control trials for the 151 index trials. These 3,330 control trials had 115,948 citations before 2025 from 87,811 unique publications, and had a median of 24 citations per publication, mean 35, range 1-375.

Figure 2 shows the mean monthly citations per publication for the index trials grouped by the type of editorial response and for the control articles. In all groups, citations increased steadily after publication, reaching their peak a median of 45-65 months after publication, before declining. As the group of controls were clearly less highly cited than the index publications, we created a highly cited control group with at least 25 citations (n=1598 control trials, 93,566 citations from 72,757 unique publications: per trial mean 59 citations, median 44, range 25-375) in case the rate or total number of citations influenced the results.

**Figure 2.**
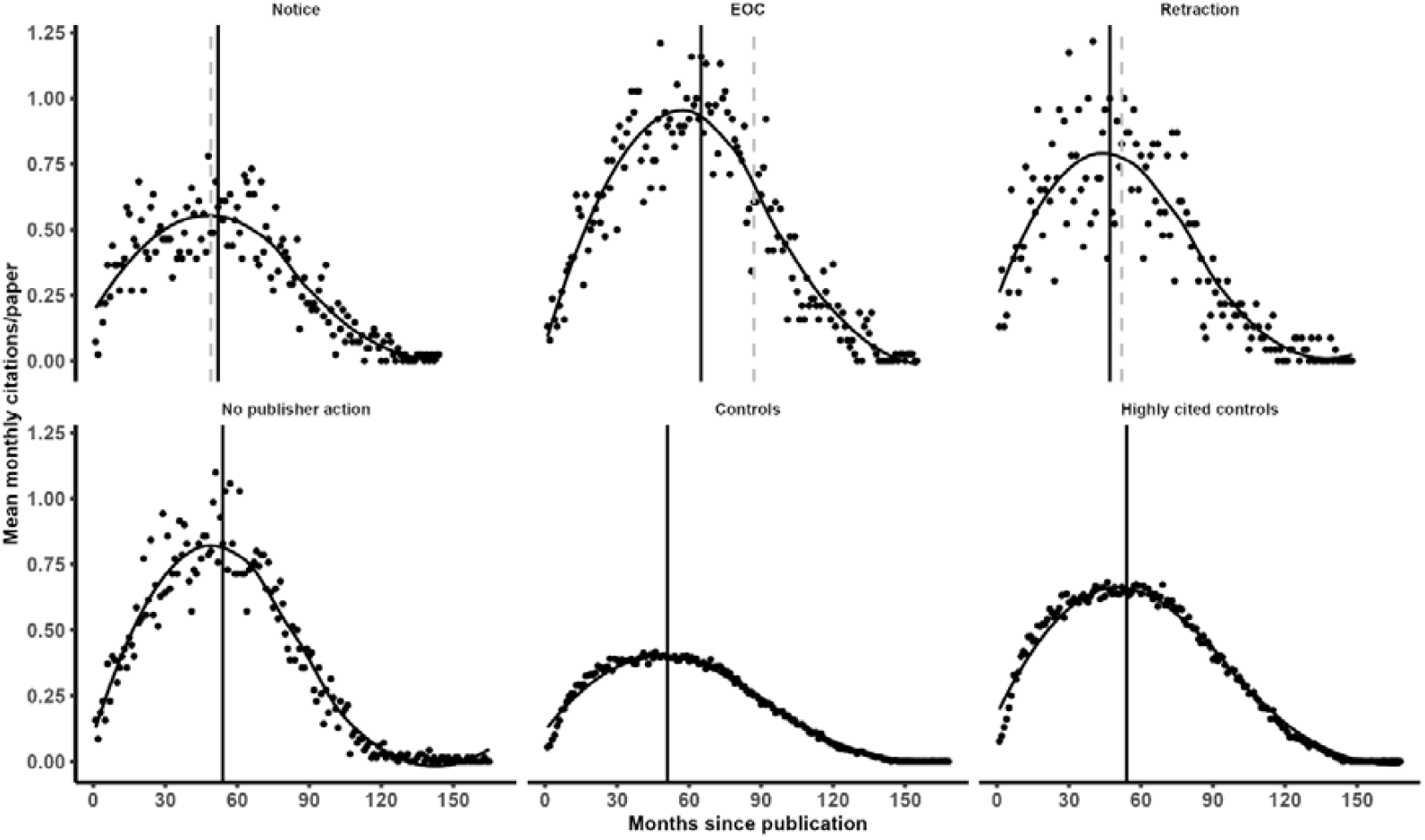
Mean monthly citations per article for the index publications grouped by the type of editorial response and for the control articles with a smoothed fit line (locally estimated scatterplot smoothing). The vertical lines are the median time to the peak citation month - the median of the months with the highest citations (solid line), and the median time to editorial response (grey dashed line).

Figure 3 shows changes in citation rates up to 60 months before and after the editorial responses for the index trials. A reference point for the articles without an editorial response and the controls is necessary to allow comparisons with citation rates before and after editorial responses. Since we expected citations to decline following editorial responses, the natural decline in citations following their peak seemed an appropriate comparator. We therefore used the peak citation month, ie the median of the months with the highest citations, as the reference point for the articles without an editorial response and the controls. Figure 3 shows that for each category of publication, changes in citation rates before and after the respective reference point were approximately linear.

**Figure 3.**
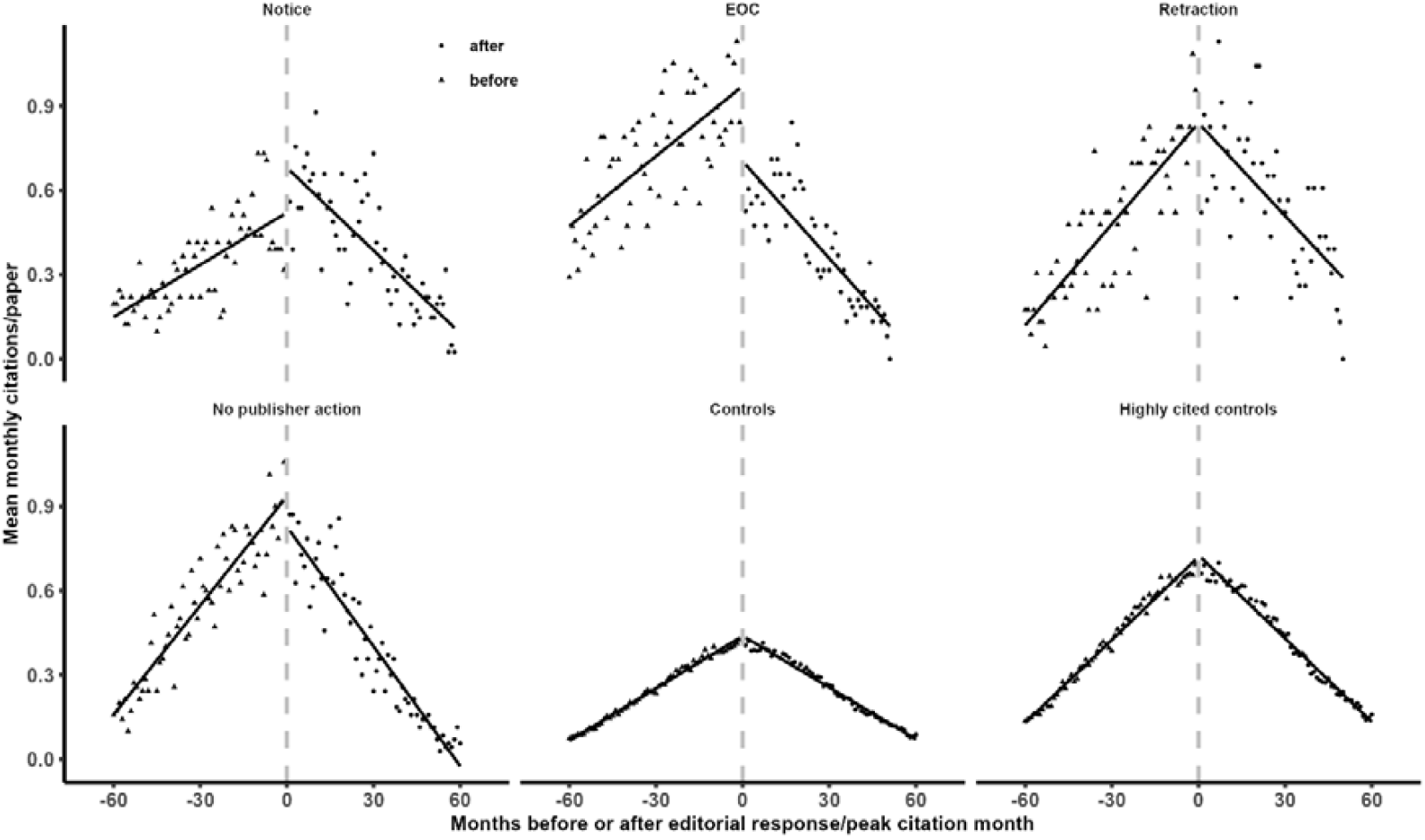
Mean monthly citations per article up to 60 months before and after the editorial response or the peak citation month for articles without an editorial response and the controls. The line is a linear regression line of best fit.

Of particular interest is whether the slopes after an editorial response differ from the natural rate of citation decline seen in the papers without editorial responses and the controls. Table 1 shows the comparison of these slopes. Before the editorial responses or peak citation month, mean monthly citations per paper increased at rate of 0.006 to 0.013 per month. For the index trials with an editorial response, mean monthly citations per paper decreased at a rate of 0.0098 to 0.012 per month after publication of the editorial response. All pairwise differences between these 3 categories were not statistically significant. For the index trials without an editorial response, the rate of decline was 0.014, higher than the rate for the trials with an EoC or a notice. The rate of decline for the controls (0.006) was lower than for all other categories but the rate of decline for the highly cited controls (0.0099) was not different in any pairwise comparison from the index trials for all 3 categories of editorial responses.

**Table 1:** Comparisons of changes in citation rates before and after editorial response or peak citation month.

| **Before** |  | **Retraction** | **EoC** | **Notice** | **No publisher action** | **Controls** | **Highly cited controls** |
| --- | --- | --- | --- | --- | --- | --- | --- |
| **Slope** |  | 0.012<br>(0.001) | 0.0083<br>(0.0011) | 0.0062<br>(0.0008) | 0.013<br>(0.00069) | 0.0062<br>(0.000083) | 0.0098<br>(0.00016) |
| **Retraction** | 0.012 |  | 0.015 | <0.001 | 0.46 | <0.001 | 0.037 |
| **EoC** | 0.0083 | 0.015 |  | 0.13 | <0.001 | 0.061 | 0.18 |
| **Notice** | 0.0062 | <0.001 | 0.13 |  | <0.001 | 0.99 | <0.001 |
| **No publisher action** | 0.013 | 0.46 | <0.001 | <0.001 |  | <0.001 | <0.001 |
| **After** |  | **Retraction** | **EoC** | **Notice** | **No publisher action** | **Controls** | **Highly cited controls** |
| **Slope** |  | -0.011<br>(0.0019) | -0.012<br>(0.0011) | -0.0098<br>(0.001) | -0.014<br>(0.00067) | -0.0061<br>(0.0001) | -0.0099<br>(0.00019) |
| **Retraction** | -0.011 |  | 0.81 | 0.55 | 0.078 | 0.0025 | 0.48 |
| **EoC** | -0.012 | 0.81 |  | 0.26 | 0.033 | <0.001 | 0.089 |
| **Notice** | -0.0098 | 0.55 | 0.26 |  | <0.001 | <0.001 | 0.95 |
| **No publisher action** | -0.014 | 0.078 | 0.033 | <0.001 |  | <0.001 | <0.001 |

The previous analyses compared the results from all the articles collectively by category. Next, we compared the slopes up to 60 months before and after the editorial response or peak citation month for each article individually. Figure 4 shows the median and interquartile range of these slopes and Table 2 shows the comparisons between the differences in the slopes before and after the editorial response or peak citation month for the categories. There were no pairwise statistically significant differences in differences in slopes amongst articles with a retraction or an EoC, those with no publisher action, and the highly cited controls (differences 0.018-0.026 citations/month per month).

**Figure 4.**
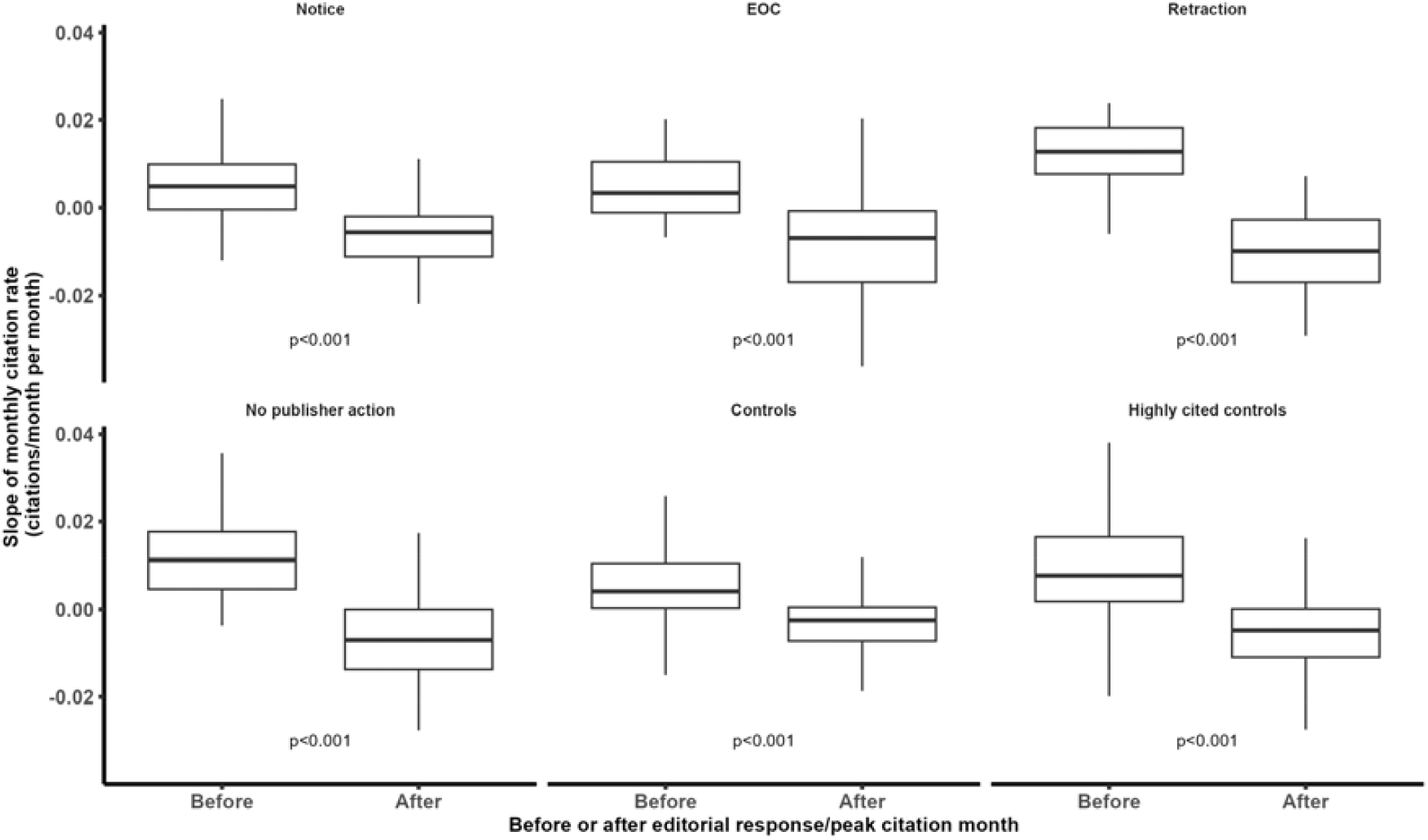
box and whisker plots of the slopes of the monthly citation rates for individual papers before and after the editorial response or peak citation month. P-values are from a Wilcoxon test.

**Table 2:** Differences between citation rates for individual publications before and after the editorial response or peak citation month.

|  |  | **Retraction** | **EoC** | **Notice** | **No publisher**<br>**action** | **Controls** | **Highly cited**<br>**controls** |
| --- | --- | --- | --- | --- | --- | --- | --- |
|  | **Difference** | 0.025<br>(0.019) | 0.026<br>(0.054) | 0.014<br>(0.013) | 0.024<br>(0.027) | 0.012<br>(0.019) | 0.018<br>(0.023) |
| **Retraction** | 0.025 |  | 0.89 | 0.024 | 0.92 | 0.0045 | 0.12 |
| **EoC** | 0.026 | 0.89 |  | 0.28 | 0.85 | 0.2 | 0.46 |
| **Notice** | 0.014 | 0.024 | 0.28 |  | 0.01 | 0.31 | 0.065 |
| **No publisher action** | 0.024 | 0.92 | 0.85 | 0.01 |  | <0.001 | 0.072 |

The difference column and row refers to the mean (SD) difference in citation rates before and after the editorial response or peak citation month for individual papers. The cells contain the p-values from a t-test of these differences between categories.

## Discussion

For this large set of 172 RCTs with integrity concerns reported by a single research group, we found little evidence that publication of an editorial response, whether it was retraction, publication of an EoC or issuing of an editorial notice, had any meaningful effect on the rate of citation. Citation rates following an editorial response in these RCTs declined at a similar rate to the natural decline in citations following their peak for contemporaneous control RCTs in the same journals with similar citation rates to the index RCTs prior to the editorial response. The median peak citation month for the index RCTs was similar to that of the control studies and very close to the median editorial response month suggesting that if publication of an editorial response led to the citation decline, it is reasonable to expect the decline to be faster than control articles without any editorial responses. As we did not observe this, it suggests that publication of an editorial response has little impact on citation rates over and above the natural decline in citations. It would also be expected that the more serious and visible editorial responses, EoC and retraction, would have greater impact than editorial notices which often have little visibility. Once again, we did not observe this, perhaps because none of the editorial responses affected citation rates. The broader conclusion is that publication of an editorial response did not meaningfully reduce subsequent references to these trials with integrity concerns. If these editorial responses do not impact upon subsequent citation rates, it raises the question of how much value they provide.

We are not aware of previous research focused on the impact of EoC or editorial notices on citations, but there is a body of research on citation rates following retraction. The findings of that research are mixed. A survey of citations to 7813 retracted articles between 1960-2020 concluded that retracted papers continue to be cited but over time citations decline.^6^ Another study reported that overall citations increased rather than decreased following retraction for 304 articles,^7^ although retracted studies in high impact journals or with many pre-retraction citations did have a decline in citations.^7^ Nearly 60% of 6926 citations to 376 articles by a single research group, were to the 113 articles with either an EoC or retraction^9^ While almost all of the citing articles were published before the editorial response, about 5% occurred afterwards and 11% of these citing articles after the editorial response referenced multiple articles with editorial responses.^9^ An analysis of 1423 retractions reported that a single retraction for misconduct led to a >80% reduction in citations to the retracted article within 5y and a 7%/year decline in citations across the whole body of the retracted author’s work.^5^ In 100 authors with multiple retractions, citation rates for retracted papers declined after the first retraction for the author but rates for unretracted papers remain similar.^10^ Collectively, these and other studies show that citations occur following retraction and may decline but almost all do not address the question of whether that decline occurs faster than unretracted control articles. One study that did address that question found a 23% reduction in citations in the month after retraction followed by a reduction of 2% per month over the next 2y relative to the expected rates in control studies.^11^

One consideration is that the editorial response may be issued too late after a paper has been published to prevent further citations. Generally, citations peak about 2-4 years after publication, although this varies considerably within and between disciplines.^12^ In our analyses, the peak citation month ranged from 48-65 months after publication (Figure 2). For the index set of trials, the editorial responses were published on average about 5y after the original publication, meaning it is possible that citation rates were already declining naturally, and an editorial notice may have had little impact on that decline. It is possible that if the editorial notice occurred earlier, the decrease in citation rates might have been greater. But it is also possible that an early retraction may simply stop any further increase in citations which then decline at the same rate as articles with similar peak citation rates. Such possibilities could be assessed in future analyses but necessarily relies upon journals and publishers taking editorial action much sooner than is currently the norm.

There are many reasons why citation of articles with published editorial responses may occur. Citations may occur before the editorial response is published. When the citation occurs after the editorial response is published, the authors may not be aware of the editorial responses for a variety of reasons: they may not check the primary source, they may copy the reference from another source, they may use a personal reference manager that does not update when a publication is retracted, or the editorial response may be difficult to find because it appears only on the publisher webpage, or is not linked to the source article in PubMed or similar search tools. For example, for 441 retracted publications on public health, only about 50% of publicly available publication records indicated that they were retracted and only 5% of articles were identified as retracted in all the 11 sources of publication records examined.^13^ Use of decentralised, non-publisher sources such as institutional repositories, personal libraries and reference management software may compound this issue.^14^ As technology improves, citation of retracted publications or those with EoC or editorial notices may become less common, as reference-management software begin to update references with changes automatically and incorporate information from retraction databases. Journals and publishers may systematically screen for such citations prior to publication. However, these improvements cannot address the issue of citations to papers that are subsequently retracted after the citing article is published. Our own experience is that few such papers are corrected, even when the retracted article provides a key rationale or is a key component of the paper.^15^

One aspect of our analysis that warrants attention is the use of the open source OpenCitations database. Detailed citation data in the form necessary to do analyses such as those in this publication can be difficult or expensive to obtain from databases run by commercial companies. The OpenCitations database offers a free alternative that when combined with the use of R and Pubmed allowed extremely detailed and sophisticated analyses of citations to occur. Meta-research may become more prevalent since the presence of large, open access databases and free statistical software allows easy access and analyses for independent researchers.

In summary, we did not find evidence that an editorial response-a retraction, an EoC, or an editorial notice-altered the rate of decline of citations above the expected natural decline. Currently, it is unclear whether existing editorial strategies effectively limit the adverse effects of unreliable publications. The new “Communication of Retractions, Removals, and Expressions of Concern” guidelines from the National Information Standards Organization^16^ may enhance the effectiveness of future editorial responses by improving their visibility and prompting more timely and consistent publisher action.

## Acknowledgements

No funding source declared. We thank Paul Manson, Information Scientist, Aberdeen Centre for Evaluation, University of Aberdeen, for initial advice on citation searching.

## Ethical approval

Not applicable.

## Guarantor

Mark Bolland

## Data availability

The data were gathered from open-access datasets that are freely available. The specific dataset can be requested from.

## Generative AI use

Generative AI was not used in the preparation of this paper.

## CRediT authorship contribution statement

Hugo Studd – conceptualization, investigation, data curation, formal analysis, writing – original draft, writing – review and editing. Alison Avenell – conceptualization, investigation, writing – review and editing. Andrew Grey – conceptualization, investigation, writing – review and editing. Mark Bolland - conceptualization, investigation, data curation, formal analysis, writing – review and editing.

For the purpose of open access, the authors have applied a Creative Commons Attribution (CC BY) [or other appropriate open licence] licence to any Author Accepted Manuscript version arising from this submission.

## References

1. Van Noorden R. More than 10,000 research papers were retracted in 2023 - a new record. Nature 2023;624:479–81.

2. Fanelli D. How many scientists fabricate and falsify research? A systematic review and meta-analysis of survey data. PLoS One 2009;4:e5738.

3. Gopalakrishna G, Ter Riet G, Vink G, Stoop I, Wicherts JM, Bouter LM. Prevalence of questionable research practices, research misconduct and their potential explanatory factors: A survey among academic researchers in The Netherlands. PLoS One 2022;17:e0263023.

4. Chen L, Li Y, Wang J, Li Y, Tan X, Guo X. Knowledge, attitudes and practices about research misconduct among medical residents in southwest China: a cross-sectional study. BMC Med Educ 2024;24:284.

5. Lu SF, Jin GZ, Uzzi B, Jones B. The retraction penalty: evidence from the Web of Science. Sci Rep 2013;3:3146.

6. Hsiao TK, Schneider J. Continued use of retracted papers: Temporal trends in citations and (lack of) awareness of retractions shown in citation contexts in biomedicine. Quant Sci Stud 2022;2:1144–69.

7. Candal-Pedreira C, Ruano-Ravina A, Fernandez E, Ramos J, Campos-Varela I, Perez-Rios M. Does retraction after misconduct have an impact on citations? A pre-post study. BMJ Glob Health 2020;5.

8. Bolland MJ, Gamble GD, Grey A, Avenell A. Empirically generated reference proportions for baseline p values from rounded summary statistics. Anaesthesia 2020;75:1685–7.

9. Bolland MJ, Grey A, Avenell A. Citation of retracted publications: A challenging problem. Account Res 2022;29:18–25.

10. Mistry V, Grey A, Bolland MJ. Publication rates after the first retraction for biomedical researchers with multiple retracted publications. Account Res 2019;26:277–87.

11. Mott A, Fairhurst C, Torgerson D. Assessing the impact of retraction on the citation of randomized controlled trial reports: an interrupted time-series analysis. J Health Serv Res Policy 2019;24:44–51.

12. Lachance C, Larivière V. On the citation lifecycle of papers with delayed recognition. Journal of Informetrics 2014;8:863–72.

13. Bakker CJ, Reardon EE, Brown SJ, et al. Identification of retracted publications and completeness of retraction notices in public health. J Clin Epidemiol 2024;173:111427.

14. Davis PM. The persistence of error: a study of retracted articles on the Internet and in personal libraries. J Med Libr Assoc 2012;100:184–9.

15. Avenell A, Bolland MJ, Gamble GD, Grey A. A randomized trial alerting authors, with or without coauthors or editors, that research they cited in systematic reviews and guidelines has been retracted. Account Res 2024;31:14–37.

16. National Information Standards Organization (NISO). Communication of Retractions, Removals, and Expressions of Concern (CREC): NISO Recommended Practice (NISO RP□45□2024); 2024.

